# CORONAVIRUS DISEASE 2019 IN A TERTIARY PEDIATRIC CENTER IN PORTUGAL

**DOI:** 10.1101/2021.08.16.21262100

**Authors:** Tiago Milheiro Silva, Ana Margarida Garcia, Catarina Gouveia, Flora Candeias, Maria João Brito

## Abstract

**Objectives:** Describe the demographic, clinical, laboratory, and imaging features of SARS-CoV-2 infected children at a tertiary pediatric center in Portugal during the first 6 months of the COVID-19 pandemic.

**Design:** Single center, descriptive study of pediatric patients, who had a confirmed SARS-CoV-2 infection from March 7 to September 20, 2020.

**Setting:** Tertiary Pediatric referral center.

**Patients:** 18 years or younger.

**Main outcome measures:** Incidence, mortality, age of infection, clinical characteristics, treatment prescribed and outcome.

**Results:** A total of 300 patients were included with a median age of 5 years (IQR 1-11) and in 67% a contact was identified (co-habitant in 52.7%). 56 (18.7%) had pre-existing medical conditions. A mode of three days mediated symptom appearance to diagnose. The most common symptoms were fever (55.7%), cough (38.3%), and nasal congestion (24%). 23% of the patients were admitted due to complications related to COVID-19 and 2% required intensive care. We used drugs with antiviral activity in 9% of the patients, immunomodulatory medication with corticosteroids in 3.3%, and intravenous immunoglobulin in 1.7%. Two (0.6%) children died and 2.3% reported short-term sequelae.

**Conclusions:** COVID-19 is usually a mild disease in children, but a small proportion of patients develop severe and critical disease. Fatal outcomes were rare overall and exclusive of severe previous medical conditions. Suspecting and diagnosing COVID-19 in children based on their symptoms without epidemiologic information and virus testing is very challenging. Our data also reflect the uncertainties regarding specific treatment options, highlighting that additional data on antiviral and immunomodulatory drugs are urgently needed.

## Introduction

Coronavirus disease 2019 (COVID-19), caused by the new severe acute respiratory syndrome coronavirus 2 (SARS-CoV-2)^1^, was first found linked to a cluster atypical pneumonia in Wuhan, China in December 2019^2^. Person to person transmission has resulted in the rapid spread of the disease across borders and continents. The World Health Organization (WHO) declared COVID-19 as a pandemic on March 11, 2020^3^, and as of September 2020, 30,675,675 cases have been confirmed and 954,417 fatal cases have been reported^4^. Europe has been one of the most affected regions^4^.

In Portugal, the first case was reported on March 2^5^ and the first pediatric case on March 7. Until September 20, there were a total of 68,577 cases^6^. Of these, 6,356 were children younger than 19 years (9.3% of total cases)^6^.

Our hospital was one of two national reference hospitals for COVID-19 pediatric patients^7^ and has been responsible for the management of a major portion of pediatric severe cases in our country.

An unprecedented amount of scientific information has been produced in a short amount of time^8^. This led to frequent changes in treatment strategies of COVID-19 patients. Hydroxychloroquine, for example, changed from being one of the most promising drugs^9^ to being removed from the *SOLIDARITY* trial^10^. Although the pediatric population seems to be less affected than adults^11^ and the clinical presentation and evolution generally less severe, critical cases occur nonetheless^12-16^. With the increasing number of new diagnoses in Europe, severe cases in children are bound to rise, including multisystem inflammatory syndrome associated with COVID-19 (MIS-C)^17^.

COVID-19 in the pediatric ages continues to be less characterized than in adults^18^. Aspects such as secondary transmission, ideal methods for diagnosis, and clinical and laboratory markers of worse prognosis are still being questioned^19^. In addition, the pulmonary imagological aspects can be different than in the adult population^20^ and therapeutic options are still limited^21^.

This study aimed to review and analyze the demographic, clinical, laboratory, and imaging features of the first SARS-CoV-2 infected patients accompanied in our hospital during the first 6 months of the pandemic.

## Methods

Single center, descriptive study of children and adolescents, 18 years or younger, who had a confirmed SARS-CoV-2 infection from March 7 to September 20, 2020. Age, sex, underlying disease, date of diagnosis, route of exposure, clinical, laboratory, and radiographic findings, treatment, and outcome were analyzed. The cases were diagnosed by real-time reverse transcription–polymerase chain reaction (rRT-PCR) in a combined nasopharyngeal and oropharyngeal swab or respiratory secretions or by the confirmation of antibodies to SARS-CoV-2.

Patients were considered to have mild, moderate, severe, or critical disease according to the WHO guidelines^22^ (Table 1).

**TABLE 1.** Clinical respiratory syndromes associated with COVID-19 according to World Health Organization (WHO).

|  |  |
| --- | --- |
| **Asymptomatic** | Without any symptoms, confirmed by laboratory tests |
| **Mild illness** | **Respiratory infection of the upper tract:** fever, cough, fatigue, anorexia, myalgia, sore throat, rhinorrhoea, headache. Rarely with nausea, vomiting and diarrhoea. |
| **Moderate case** | **Pneumonia with non-severe signs of respiratory distress** (tachypnoea < 2 months: $\geq 60$ cpm; 2–11 months: $\geq 50$ cpm; 1–5 years: $\geq 40$ cpm, >5 years: >30 cpm, moderate recession), $SpO_2 \geq 93\%$ |
| **Severe case** | **Pneumonia with severe signs of respiratory distress** (central cyanosis or $SpO_2 < 93\%$, severe tachypnoea (<12 months: >70 cpm; >12months: >50 cpm), severe recession, grunting, nasal flaring), severe anorexia, lethargy/diminished level of consciousness, convulsions |
| **Critical cases** | Acute respiratory distress syndrome (ARDS), sepsis, septic chock |

ARDS - Acute respiratory distress syndrome; cpm – cycles per minute; $SpO_2$ - peripheral capillary oxygen saturation

All antiviral therapies were administered after informed consent was signed by parents or caretakers and approval by the pharmacy commission of our hospital was obtained.

The differences between the groups were assessed through univariable analysis using the chi-square test or Fisher exact probability test for categorical variables. P-values of <0.05 were considered statistically significant. Odds ratio (OR) and 95-Confidence Intervals were calculated to estimate how each factor influenced the assessed risks. All of the analyses were conducted in Stata^®^ version 14.1.

No Patients or Public were involved in the execution of this study.

## Results

Three hundred patients were included. May was the month with a higher count of new diagnoses (27%), accompanying, with some delay, the Portuguese epidemiologic curve (Figure 1). The median age was 5 years (IQR 1-11), 24.3% were younger than 1 year, and 4% younger than one month, with a male to female ratio of 1.11 (Table 2). A known contact was identified in 67% patients. The most common source of infection was a household contact (52.7%): 41% had at least one parent infected and 11.7% another adult co-habitant. In 4.3%, infection was nosocomial.

**FIGURE 1.**
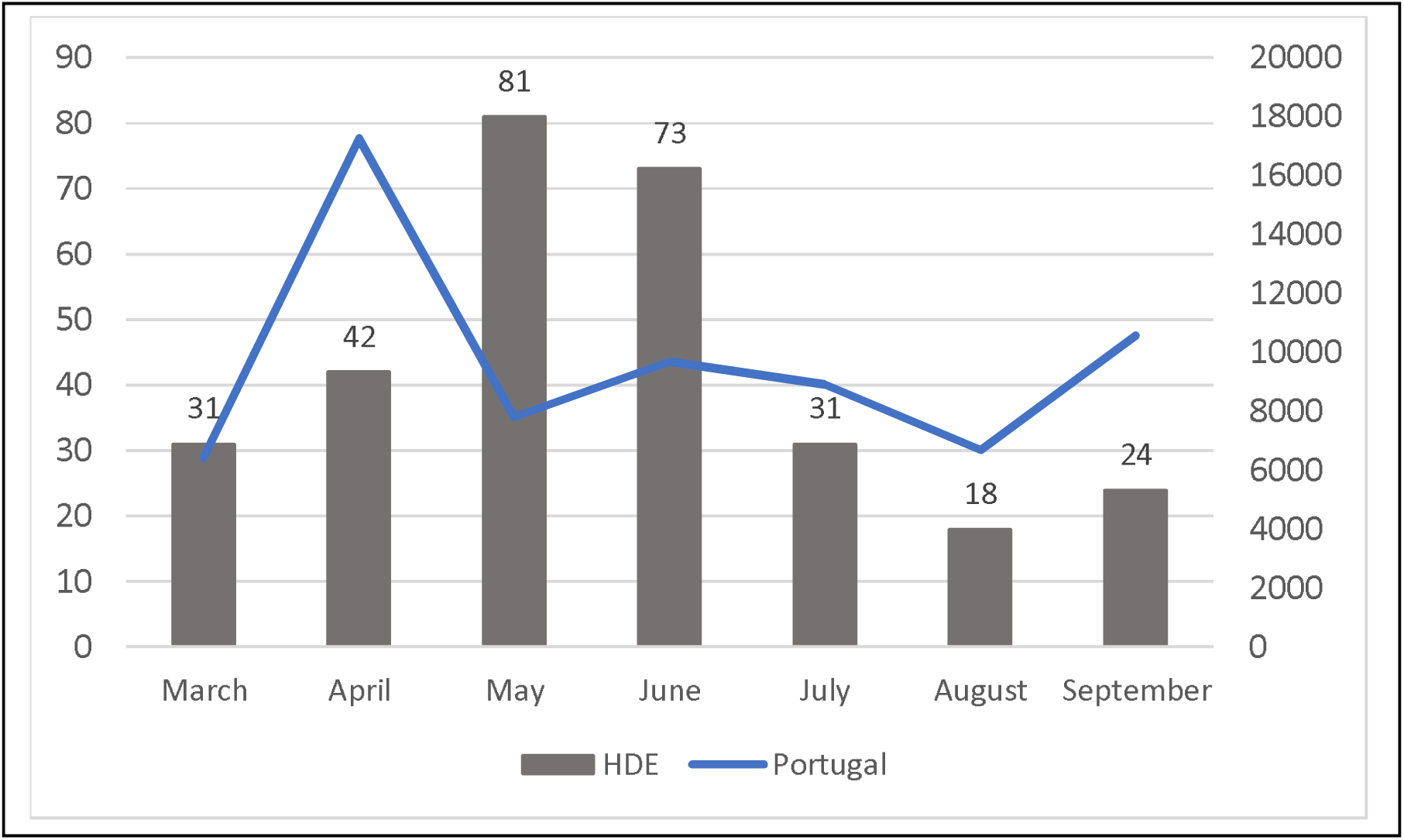
Monthly distribution of new diagnosis in our hospital and new diagnosis in the Portuguese population

**TABLE 2.**
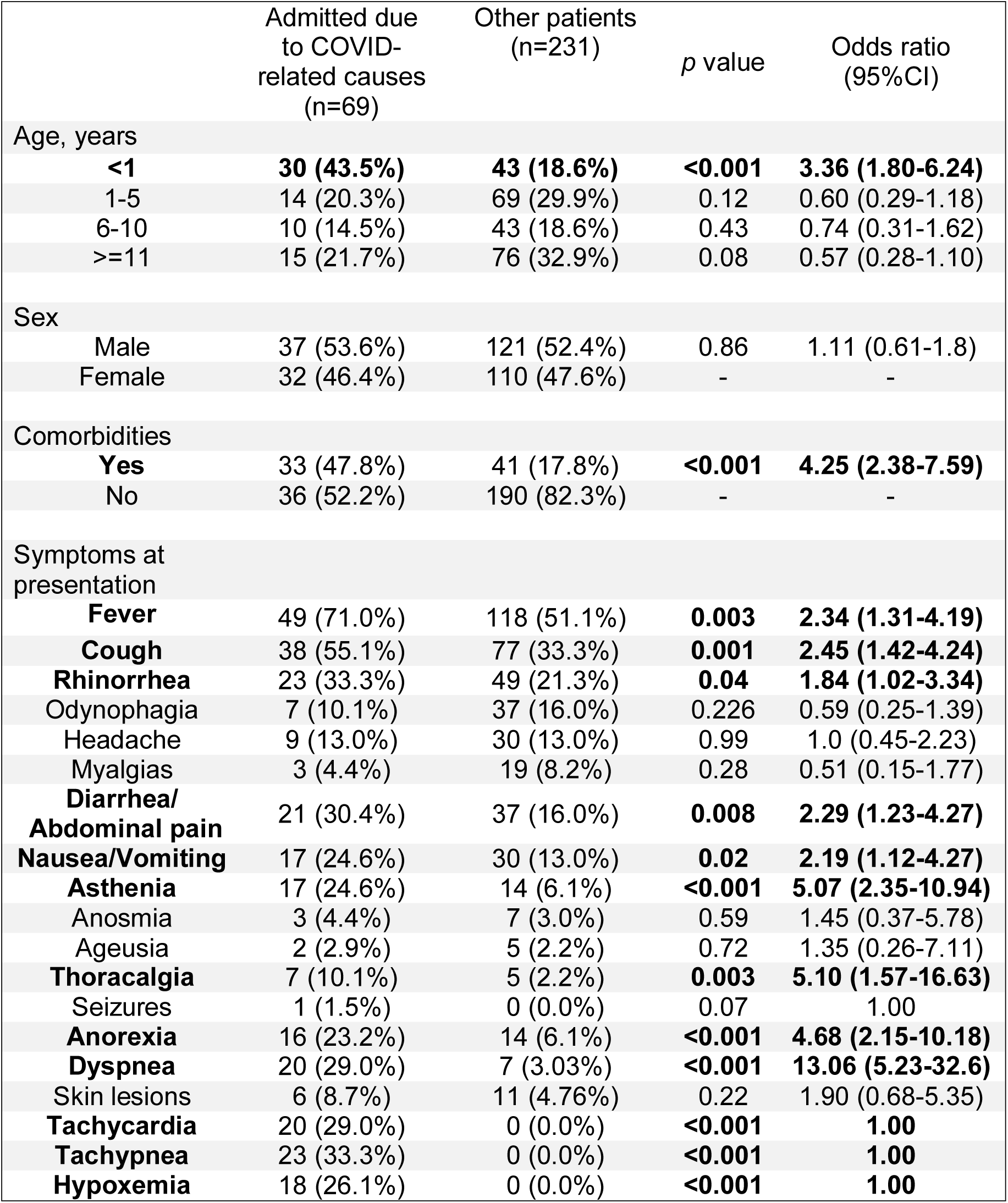
Demographic and clinical characteristics in admission.

|  | Admitted due to COVID-related causes (n=69) | Other patients (n=231) | p value | Odds ratio (95%CI) |
| --- | --- | --- | --- | --- |
| Age, years |  |  |  |  |
| <1 | **30 (43.5%)** | **43 (18.6%)** | **<0.001** | **3.36 (1.80-6.24)** |
| 1-5 | 14 (20.3%) | 69 (29.9%) | 0.12 | 0.60 (0.29-1.18) |
| 6-10 | 10 (14.5%) | 43 (18.6%) | 0.43 | 0.74 (0.31-1.62) |
| >=11 | 15 (21.7%) | 76 (32.9%) | 0.08 | 0.57 (0.28-1.10) |
| Sex |  |  |  |  |
| Male | 37 (53.6%) | 121 (52.4%) | 0.86 | 1.11 (0.61-1.8) |
| Female | 32 (46.4%) | 110 (47.6%) | - | - |
| Comorbidities |  |  |  |  |
| **Yes** | 33 (47.8%) | 41 (17.8%) | **<0.001** | **4.25 (2.38-7.59)** |
| No | 36 (52.2%) | 190 (82.3%) | - | - |
| Symptoms at presentation |  |  |  |  |
| **Fever** | 49 (71.0%) | 118 (51.1%) | **0.003** | **2.34 (1.31-4.19)** |
| **Cough** | 38 (55.1%) | 77 (33.3%) | **0.001** | **2.45 (1.42-4.24)** |
| **Rhinorrhea** | 23 (33.3%) | 49 (21.3%) | **0.04** | **1.84 (1.02-3.34)** |
| Odynophagia | 7 (10.1%) | 37 (16.0%) | 0.226 | 0.59 (0.25-1.39) |
| Headache | 9 (13.0%) | 30 (13.0%) | 0.99 | 1.0 (0.45-2.23) |
| Myalgias | 3 (4.4%) | 19 (8.2%) | 0.28 | 0.51 (0.15-1.77) |
| **Diarrhea/ Abdominal pain** | 21 (30.4%) | 37 (16.0%) | **0.008** | **2.29 (1.23-4.27)** |
| **Nausea/Vomiting** | 17 (24.6%) | 30 (13.0%) | **0.02** | **2.19 (1.12-4.27)** |
| **Asthenia** | 17 (24.6%) | 14 (6.1%) | **<0.001** | **5.07 (2.35-10.94)** |
| Anosmia | 3 (4.4%) | 7 (3.0%) | 0.59 | 1.45 (0.37-5.78) |
| Ageusia | 2 (2.9%) | 5 (2.2%) | 0.72 | 1.35 (0.26-7.11) |
| **Thoracalgia** | 7 (10.1%) | 5 (2.2%) | **0.003** | **5.10 (1.57-16.63)** |
| Seizures | 1 (1.5%) | 0 (0.0%) | 0.07 | 1.00 |
| **Anorexia** | 16 (23.2%) | 14 (6.1%) | **<0.001** | **4.68 (2.15-10.18)** |
| **Dyspnea** | 20 (29.0%) | 7 (3.03%) | **<0.001** | **13.06 (5.23-32.6)** |
| Skin lesions | 6 (8.7%) | 11 (4.76%) | 0.22 | 1.90 (0.68-5.35) |
| **Tachycardia** | 20 (29.0%) | 0 (0.0%) | **<0.001** | **1.00** |
| **Tachypnea** | 23 (33.3%) | 0 (0.0%) | **<0.001** | **1.00** |
| **Hypoxemia** | 18 (26.1%) | 0 (0.0%) | **<0.001** | **1.00** |

Fifty-six (18.7%) patients had a pre-existing chronic medical condition: chronic pulmonary disease (18), congenital heart disease (9), chronic neurologic disease (7), chromosomal abnormalities (5), chronic hematological disease (4), short bowel syndrome (3), chronic renal disease with end stage kidney failure (3) and type 1 diabetes (2). Four patients were receiving immunosuppressive therapy.

Among the 300 children, 57 (19%) were asymptomatic. Demographic and clinical characteristics of patients are showed in table 2.

Fever was more common in children below 5 years of age. Dyspnea, tachycardia, tachypnea, and hypoxemia were more frequent in younger children (<1year), while odynophagia, headache, myalgia, anosmia, ageusia, and thoracalgia were more frequent in older children and adolescents. Cough and rhinorrhea occurred equally in all age groups. Thirty-five patients (11.7%) with gastrointestinal symptoms presented no respiratory symptoms, although the majority (20/35) also presented fever.

The mode interval of the time between symptom appearance and diagnosis was three days (IQR 1-4).

One hundred and seventeen (39%) patients were admitted, 69 (59%) with COVID-19 and 48 (41%) for other reasons, namely diagnosis during the containment phase (6,8%), social motives (1.7%), and other conditions requiring hospitalization not related to COVID-19 (32.5%) such as surgical conditions, infections with need of IV antibiotics and psychiatric conditions. Admitted patients with COVID-19 were classified as having a mild disease in 27 cases (39.1%), moderate in 21 (30.4%), severe in 10 (14.5%), and critical in 11 (15.9%) (Table 3).

**TABLE 3.** COVID-19 disease classification and diagnosis in admitted patients.

| **COVID19 disease Classification and diagnosis** | **N (%)** |
| --- | --- |
| **Mild cases** | **27 (39,1)** |
| Mild illness without comorbidities | 16 (23.2) |
| Mild illness with comorbidities | 11 (15.9) |
| **Moderate cases** | **21 (30.4)** |
| Pneumonia | 21 (30.4) |
| **Severe cases** | **10 (14.5)** |
| Pneumonia | 10 (14.5) |
| **Critical cases** | **11 (15.9)** |
| MIS-C | 4 (5.8) |
| Pneumonia | 1 (1.4) |
| ARDS | 2 (2.9) |
| Myocarditis | 2 (2.9) |
| Sepsis | 2 (2.9) |

In children admitted with COVID-19, the most frequent findings were leucocytosis (47.8%), lymphocytosis (47.8%), LDH (72.9%), and elevated inflammatory markers such as procalcitonin (71.9%), ferritin (56.9%), and CRP (21.7%). Lymphopenia was present in 13%.

A chest X-ray was performed in all of the admitted patients, 59.4% with abnormal findings and a chest CT was performed in 31/69 (44.9%) patients, with abnormal findings in 25/31 (80.6%). According to the *British Society of Thoracic Imaging codes*^23^, we found classic COVID-19 findings with bilateral ground glass opacities (11), peripheral consolidations (8), reverse halo (1), and a perilobular pattern (1). Nine patients had findings indeterminate with unilateral ground glass opacities and three patients had findings non-typical for COVID-19 [lobar pneumonia (2), pleural effusion (2), and lymphadenopathy (1)].

Respiratory co-infection occurred in 27.5% of admitted patients with rhinovirus (9), adenovirus (3), enterovirus (3), influenza B (2), and *Pneumocystis jiroveci* (2). Other respiratory pathogens included syncytial respiratory virus, metapneumovirus, coronavirus 43, parainfluenza 3, bocavirus, and *Mycoplasma pneumoniae*.

Six patients were admitted to the PICU with an average length of stay of 10 days: MIS-C patients who needed inotropic support and mechanical ventilation (2), myocarditis in patients with known cardiopathy (2), COVID-19 pneumonia and ARDS in a patient with lymphangioendotheliomatosis, and another patient with chronic respiratory insufficiency.

Antiviral therapy was proposed for 21 (20.4%) patients, including mild disease with comorbidities (3) moderate (5), severe (6), and critical (7) disease. Hydroxychloroquine (HCQ) was administered in 13 (18.8%) patients, in monotherapy (8) or combined with lopinavir/ritonavir (LPV/r) (5). LPV/r was administered in four patients in monotherapy. Five (7.2%) patients were treated with remdesivir. Children admitted due to mild illness without risk factors (23.2%) did not receive therapy. In mild disease with preexisting medical conditions (15.9%), therapy was decided on a case-by-case basis. Regarding moderate disease (30.4%), sixteen patients (21.7%) did not receive antiviral treatment due to favorable clinical evolution. The other cases of pneumonia (5) were treated with HCQ (1), LPV/r (2), and HCQ combined with LPV/r (2). Patients with hypoxemic pneumonia with severe signs of respiratory distress (10; 14.5%) were treated with HCQ (3), and HCQ combined with LPV/r (3). Four patients with severe disease did not receive treatment according to clinical evolution. Critical cases (11) included sepsis in patients with less than three months of age treated with LPV/r (2) and one patient with ARDS received HCQ. Remdesivir was administered in three patients with myocarditis and one in a child with ARDS. Four patients with MIS-C and one case of severe ARDS were treated with intravenous immunoglobulin and systemic corticosteroids. Systemic corticosteroids were still administered in the exacerbation of asthma (4) and one patient with chronic respiratory insufficiency.

Antibiotics were used in 30 (43.5%) patients. Ceftriaxone, amoxicillin with clavulanate, cefuroxime, and/or clindamycin when respiratory bacterial superinfection was suspected (17), ampicillin, cefotaxime, and gentamycin in infants less than 3 months old (2), and vancomycin, ceftriaxone, and clindamycin empirically in the admission of MIS-C (4). Two patients with *Pneumocystis jiroveci* coinfection received cotrimoxazole. Other treatments with azithromycin and oseltamivir were used when co-infections by *Mycoplasma pneumonia* and influenza were suspected.

27.5% of admitted patients needed supplemental oxygen, for a mean duration of 6.9 days (range 1-38) and mechanical ventilation was needed in four patients with a mean duration of 10 days (range 5–20). Inhaled therapy with bronchodilators and corticosteroids was used in 57.1% of patients. Prophylactic enoxaparin was used in three patients with MIS-C and in one patient with sickle cell disease and acute chest syndrome.

Adverse drug reactions were seen in three patients who received LPV/r with nausea and vomiting, but with no need to discontinue therapy. Among the 13 patients who received HCQ, regular electrocardiograms were undertaken and only two had to discontinue HCQ temporarily (24 hours) due to prolonged QT interval (QTc) > 500 msec. No adverse effects were seen with remdesivir.

The mean hospital duration of stay was 6.8 days, ranging from 1 day to 28 days.

Two patients (0.7%) had a fatal outcome, both with severe comorbidities (neurologic disease and congenital cardiopathy).

The mean duration time between the first positive and the first negative rRT-PCR was 23 days, with a range of 1 and 115 days.

Seven children (2.3%) reported sequelae: increased oxygen therapy in an infant with bronchopulmonary dysplasia (1); myocardial scar tissue in cardiac magnetic resonance (1), worsening of cardiac function in a child with severe aortic valve regurgitation (1), and weight loss and muscular atrophy in patients with MIS-C (4). A child with dilated cardiomyopathy and myocarditis showed worsening of myocardial function, thereby requiring heart transplantation.

## Discussion

The COVID-19 pandemic has placed a high burden on the medical community^24^.

To our knowledge, this is the largest series of pediatric patients in Portugal, and one of the largest in Europe. We are a tertiary, referral center for pediatric patients and, as such, the study population represents individuals at the more severe disease spectrum.

Children at any age are at risk of being infected^12-17,25,26^. In our population, the median age was 5 years, in concordance with the ptbnet group study of European pediatric patients^15^, although lower than the CDC MMWR (11 years)^13^ and Dong *et al*. in China (10 years)^12^ and higher than the CONFIDENCE study (3.3 years)^25^.

In our study, the most common source of infection was a household member (52.7%) in concordance with other published data^15,25^. These numbers are lower than large series from China and the United States, 76.6^16^ and 91.3%^13^, respectively. In our sample, there was also a significant number of children with unknown exposition (33%). This brings to light the potential role of asymptomatic, non-diagnosed patients in the transmission of the virus in the community^27^, outside the house environment, and the need for the mass testing of the general population^28^. The rates of asymptomatic children have varied from 4.4-23%^12,13,15-16,25^ and probably represent a significant underestimation as many asymptomatic children are not screened. In our population, these asymptomatic patients represent co-habitants of infected people and patients who were diagnosed during hospital pre-admission screening or during control of nosocomial transmission.

The most common presentation in our population was fever (55.7%), cough (38.3%), and coryza (24%) unlike other studies^12,13,15,16^

Anosmia, a common symptom in adults^29^, has been referred to in 3.6% of our patients. Probably this represents an underreport since anosmia is difficult to elicit in younger children^30^. Patients with gastrointestinal signs and no respiratory symptoms represented 11.7%. This points to the fact that children with no respiratory symptoms must also be screened^15^. The mean interval of time between symptom appearance and diagnosis was 3.2 days. This is similar to the other published pediatric series^31^.

We had a hospital admission rate of 39%. Patients younger than 1 year, presenting with respiratory distress signs and those with pre-existing medical conditions, were significantly more likely to be admitted. The high admission rate reported relates to the fact that we function as a referral center for the entire south area of the country.

Respiratory viral coinfection occurred in 16.2% of the COVID-19 admitted patients. This is a lower number than what has been previously reported^32^. The increased use of masks and the quarantine measures instituted (e.g. closing of schools) probably explains this discrepancy^33^.

In children with SARS-CoV-2, treatment primarily consists of supportive care, including oxygen and advanced respiratory support, hydration, nutritional support, and antipyretics^34^. A various number of drugs have been proposed as treatment options in children and their use has been reported and encouraged by the national guidelines^15^. Some of them have, since the beginning of the pandemic, been withdrawn from regular clinical use^10^. Examples are lopinavir/ritonavir, which was proposed to act through the inhibition of SARS-CoV-2 proteinases^35^. It was stopped due to the lack of proven efficacy and concerns regarding its pharmacodynamics^10^, and hydroxychloroquine, proposed for the inhibition of SARS-CoV-2 entry into cells and host immunomodulatory effects^36^, which was stopped due to the lack of proven efficacy and the concern about the relevant side effects (QTc prolonging effect, arrythmias)^37^. Remdesivir has emerged as the preferred agent for treating severe COVID-19 in children^33,38^. Our data reflects the uncertainties regarding drug treatment options for COVID-19 since the beginning of the pandemic.

Immunomodulatory therapy is reserved for patients with evidence of hyperinflammation^39^. In addition, patients with known asthma or airway hyperreactivity should continue their inhaled corticosteroid and, in case of exacerbation, systemic steroids should be considered.

Infection of the respiratory system and endothelial cells by SARS-CoV-2 causes an intense inflammatory response that includes the activation of the hemostatic system^40^. As such, enoxaparin should be considered in patients with MIS-C and/or patients with other known risk factors for thrombosis^41^.

Rare pediatric fatal cases have been reported^15^. In our study, both fatal cases had severe pre-existing medical conditions.

In our study, seven children (2.3%) reported sequelae. However, these are probably underrated. It does not include, for example, psychological sequelae. Exercise intolerance, reduced lung vital capacity, myocardial scars in MIS-C patients, and loss of muscle mass are just some of the possible sequelae that need to be identified as soon as possible^42^.

## Conclusions

Our findings add to the general feeling that COVID-19 is usually a mild disease in children, but a small proportion of patients develop severe and critical disease.

Suspecting and diagnosing COVID-19 in children based on their symptoms without epidemiologic information and virus testing is very challenging. A significant proportion of patients is likely to be asymptomatic and the only way of diagnosing those patients is mass testing in the community.

Uncertainty still remains regarding specific antiviral options as more robust studies are bound to arrive in the near future.

We hope our study clearly details our experience with these patients and adds to the general information of this global health problem.

### What is known about this topic

- COVID-19 is usually a mild disease in children, but a small proportion of patients develop severe and critical disease.
- Suspecting and diagnosing COVID-19 in children based on their symptoms without epidemiologic information and virus testing is very challenging.
- COVID-19 in pediatric ages continues to be less characterized than in adults.

### What this study adds

- Suspecting and diagnosing COVID-19 in children based on their symptoms without epidemiologic information and virus testing is very challenging.
- A significant proportion of patients is likely to be asymptomatic and the only way of diagnosing those patients is mass testing.
- Our data reflects the uncertainties regarding drug treatment options for COVID-19 since the beginning of the pandemic.

## Data Availability

No additional data is available

